# Longitudinal improvement in postural instability is distinct from gait in treated adult hydrocephalus

**DOI:** 10.1101/2022.12.05.22282985

**Authors:** Amber Wacek, James Jean, Alec Jonason, Jacob T. Hanson, Vibha Mavanji, Elise F. Palzer, Scott Lewis, James Ashe, John M. Looft, Robert A. McGovern

## Abstract

**Background:** Ventriculoperitoneal shunting improves gait in patients with normal pressure hydrocephalus. Postural instability is a major concern, but mostly ignored in the evaluation and treatment of these patients. This study quantified postural instability using kinematics via a prospective cohort design.

**Methods:** Seventeen patients with suspected normal pressure hydrocephalus and twenty age-matched, healthy controls underwent quantitative pull test and gait examinations while wearing inertial measurement units at baseline. Patients with suspected normal pressure hydrocephalus who were shunted (n=13) and not shunted (n=4) underwent further testing after a lumbar drain trial and at follow-up visits 6 and 12 months post-operatively.

**Results:** While most gait improvement in patients who were shunted was seen immediately after the lumbar drain trial, measures of their postural response continued to improve after the lumbar drain trial through one year of follow-up. Patients who were not shunted showed no statistically significant changes in gait and postural instability measures.

**Conclusions:** After shunting, postural instability improves continuously over one year. In contrast, a large improvement in gait is seen immediately with minimal change over the subsequent year. This difference in timing may implicate two distinct neurophysiological mechanisms of recovery and provides novel evidence that postural instability improves in response to long-term CSF diversion.

## INTRODUCTION

Normal pressure hydrocephalus (NPH) is a disease affecting gait, balance, cognition and urinary continence that is primarily seen in people after their sixth decade of life.^1^ Gait and balance dysfunction are the most common symptoms seen in NPH. Ventriculoperitoneal shunting (VPS) has been shown to improve some gait parameters in patients with NPH for up to ten years post-operatively.^2-5^ Balance dysfunction, or postural instability (PI), is another primary concern of patients with NPH, as they frequently present with falls, but little is known about the pathophysiology of PI or its response to VPS.

Our previous work has demonstrated that the clinical gold standard assessment of PI - the pull test (Movement Disorder Society-Unified Parkinson’s Disease Rating Scale item 3.12; MDS-UPDRS_PT_) - can be instrumented and quantified with inertial measurement units (IMUs) in these patients.^6^ The goal of this study was to quantify the long-term effects of VPS on gait and PI compared to patients who did not undergo VPS using prospective kinematics collected during clinical visits.

## MATERIALS AND METHODS

### Subjects

Seventeen patients being clinically evaluated for NPH and 20 age-matched healthy control participants were prospectively enrolled over a period of two years from the Minneapolis VA Health Care System (MVAHCS) and University of Minnesota (UMN). Potential control participants with a positive medical screen for movement, gait, or balance disorders were excluded. Patients with possible NPH were excluded if unable to give consent. Demographic information was also collected (Supplemental Table 1).

Patients with suspected NPH underwent kinematic, neuropsychological, and physical therapy assessments before and after a lumbar drain trial (LDT). The LDT was completed via cerebrospinal fluid (CSF) drainage for three days and patients were re-assessed after removal on the morning of the 4th day. The treating neurosurgeon used these assessments to decide whether to offer treatment with VPS placement.

The surgical decision was mainly based on improvements in gait velocity as assessed by physical therapy and IMU-instrumented clinical gait examination. However, other factors such as cognitive improvements and consultation with the patient and family could influence the ultimate decision to offer surgery. Regardless of the surgical decision, all patients were prospectively followed with continued assessments over 12 months, relative to either lumbar drain trial date or surgical date respectively. Healthy controls were seen for baseline kinematic assessments only. This study was approved by the MVAHCS and UMN Institutional Review Boards, and all participants provided informed consent for participation according to the Declaration of Helsinki.

### Task Details

Body segment lengths were measured for each patient at their baseline testing session. Participants were then instrumented with 15 IMUs (Xsens, Enschede, Netherlands) recording at 60 Hz during each kinematic assessment session (Supplemental Fig. 1). Participants walked at their comfortable walking speed for approximately 10 meters for one or two gait assessment trials, depending on physical ability.

PI was assessed by 10-20 pull tests for each patient by a trained clinical examiner (R.M.), based on the instructions from the MDS-UPDRS_PT_.^7^ Each pull test consists of a retropulsive perturbation at the patient’s shoulders inducing a reactionary stepping response to regain postural stability. In order to fully assess the entire spectrum of balance dysfunction, clinical discretion was used to unpredictably vary the pull force intensities throughout the trials.^6^ However, the patient’s pull test was scored solely on the first trial after the instructional trial, which was always “brisk,” as required by the MDS-UPDRS_PT_ scale.^7^

### Data Collection and Variable Definition

Three-dimensional biomechanical models were created for each participant based on limb segment measurements as well as IMU sensor placement and orientation.^8^ Igor Pro 6.00 (Wavemetrics, Oregon, USA) was used to calculate center of mass and foot position, velocity, and acceleration from the recorded IMU data. Custom functions were then used to identify the relevant points of interest (Supplemental Methods, Supplemental Fig. 2). Relevant data points were verified via timestamps cross-referenced in Visual 3D (C-Motion Inc, Germantown, MD). The relevant data from each trial were then exported into R (v 3.6.2) and analyzed.

The outcome of a participant’s reactive postural response to each pull test is characterized by the participant’s COM velocity (V_COM_). A “normal” pull test response is typically characterized by a sharp rise to peak V_COM_ as the participant is briefly pulled backwards, followed by an abrupt decrease in V_COM_ as the participant recovers quickly within two steps (Figure 1A, Controls). Conversely, “abnormal” pull test responses are typically characterized by slower rises to peak V_COM_ as the patient reacts more slowly, followed by slower decreases to zero as the patient takes more steps to recover (Figure 1A, Patients).^6^ Pull test step length (PTSL) was defined as the difference in foot position between the first step onset and first step land, and reaction time was defined as the time between the perturbation onset and the initiation of the first step (Supplemental Fig. 2). When varying the pull intensity over a series of pulls, a “normal” recovery is characterized by the ability to appropriately scale initial step length and reaction time to perturbation intensity.^6^ Additional details on variable definitions can be found in our supplemental material.

**Figure 1.**
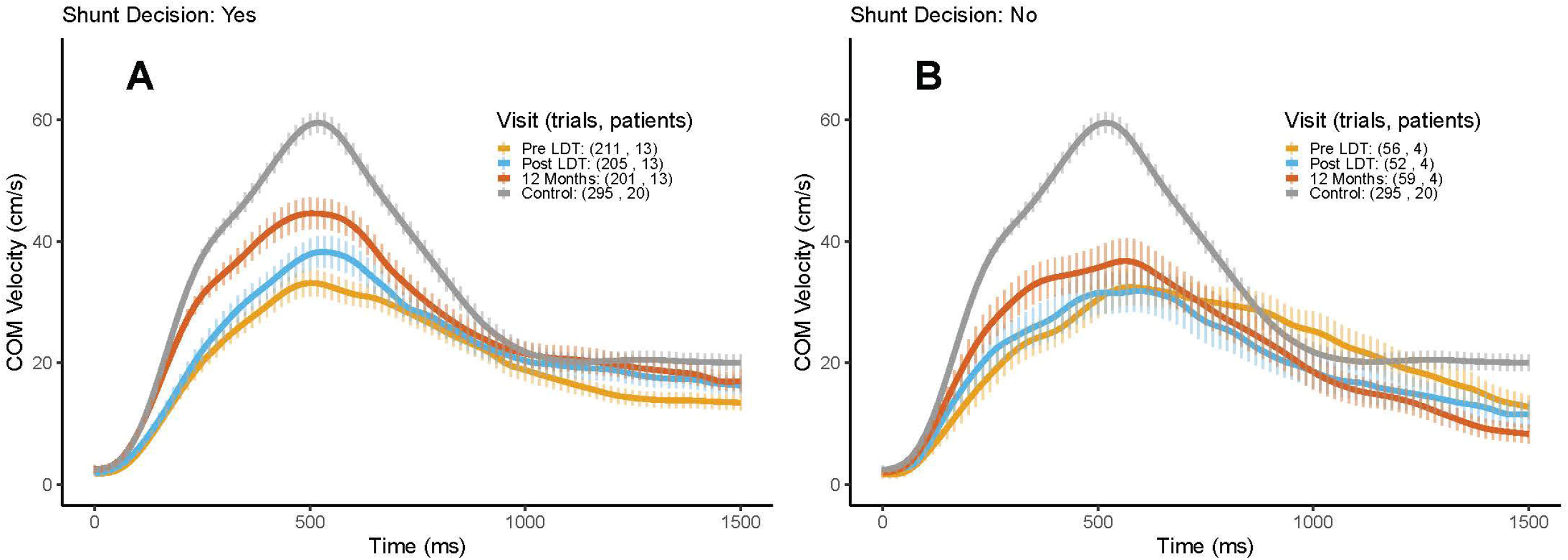

### Statistical Methods

Pull test responses for each participant were grouped by visit and VPS status. Kinematic variables and MDS-UPDRS scores between shunt decision groups were summarized by mean and standard deviation (SD) at each visit and analyzed by linear models adjusting for baseline value. Gait step count was additionally adjusted for total distance. Within-patient improvements between visits were assessed by absolute change from baseline. Between-patient improvements were not assessed due to large variability.

Mixed-effects models were used to test for within-patient changes in pull test kinematic parameters over time. These models adjusted for pull intensity, characterized as the peak value of the COM acceleration backwards (A_COM_) prior to a participant taking a step, since prior work has shown its strong association with pull test measures.^6^ Mixed-effects models additionally adjusted for baseline value. Gait and stride length changes over time were assessed by linear models adjusted for baseline value. All models used a Bonferroni p-value correction to account for multiple testing.

Mean and 95% confidence intervals (CIs) for V_COM_ were graphed over time. Other measures were graphed using mean and 95% CI for each visit. VPS status and pull test measures were plotted against A_COM_ with linear trend lines to assess potential trends. All asterisks on figures denote statistically significant changes when compared against all other visits based on the linear or mixed-effects model results after adjustment and p-value correction. All graphs and analyses were conducted using R (v 3.6.2).

### Data availability

The data that support the findings of this study are available from the corresponding author, upon reasonable request.

## RESULTS

### Improvement in the reactive postural response over time in NPH patients undergoing VPS

Treatment groups were of comparable age (Supplemental Table 1) and baseline UPDRS gait and pull test scores (Table 1). Control participants had similar ages to treatment groups but as expected, exhibited lower UPDRS scores (Table 1).

**Table 1:** Gait variables and UPDRS scores between groups over time.

| Variable<br>Mean<br>(SD) | Gait Stride Velocity (m/s) |  |  |  | Gait Stride Length (m) |  |  |  | Gait Step Count |  |  |  |
| --- | --- | --- | --- | --- | --- | --- | --- | --- | --- | --- | --- | --- |
| *Shunt Decision* | Yes | No | Control | P-value* | Yes | No | Control | P-value* | Yes | No | Control | P-value* |
| *Baseline* | 0.48 | 0.39 | 1.13 | 0.388 | 0.62 (0.23) | 0.46 | 1.21 | **0.001** | 38.61 | 48.00 | 14.5 | 0.417 |
| *Post LDT* | 0.74 | 0.37 | n/a | **<0.001** | 0.87 (0.22) | 0.46 | n/a | 0.245 | 33.38 | 44.75 | n/a | 0.169 |
| **6** | 0.75(0.17) | 0.51 | n/a | 0.066 | 0.92 (0.28) | 0.57 | n/a | **0.001** | 25.32 | 47.00 | n/a | **0.004** |
| **12** | 0.83 | 0.41 | n/a | **0.001** | 0.95 (0.22) | 0.49 | n/a | **<0.001** | 28.69 | 50.75 | n/a | **<0.001** |

| Variable<br>Mean<br>(SD) | UPDRS Gait Score |  |  |  | UPDRS Pull Test |  |  |  |
| --- | --- | --- | --- | --- | --- | --- | --- | --- |
| *Shunt Decision* | Yes | No | Control | p-value* | Yes | No | Control | P-value* |
| *Baseline* | 2.23 | 2.00 | 0.00 | 0.662 | 1.69 (0.63) | 1.75 | 0.30 | 0.889 |
| *Post LDT* | 1.62 | 1.75 | n/a | 0.252 | 1.46 (0.66) | 1.75 | n/a | 0.133 |
| 6 | 1.08 | 2.25 | n/a | **0.001** | 0.69 (0.75) | 2.25 | n/a | 0.061 |
| 12 | 1.00 | 2.5 (0.58) | n/a | **<0.001** | 0.92 (0.95) | 2.00 | n/a | **0.006** |

\*p-values are calculated between Shunt Decision groups across each visit using Wilcoxon tests, $p \leq 0.05$ are bolded.

Figure 1A depicts the improved reactive postural response of NPH patients undergoing VPS across visits. After adjusting for pull intensity, statistically significant increases in peak V_COM_ were found from baseline to post-LDT (*p*<0.001, 95% CI [3.16, 6.17]) and to one-year post-VPS (*p*<0.001, 95% CI [7.18, 10.86]). Shorter time to peak V_COM_ from baseline to one-year post-VPS (*p*=0.00, 95% CI [-0.12, -0.04]) were also observed (Supplemental Table 2). Non-shunted patients demonstrated no significant improvement in postural responses as measured by peak V_COM_ over the 12-month follow-up period (Figure 1B, Supplemental Table 2).

Due to sex differences between NPH (88.2% male) and control groups (20.0% male), we graphed pull test responses between men and women and found no visible difference in peak V_COM_ between men and women, regardless of MDS-UPDRS_PT_ score (Supplemental Fig. 3).

### Gait and postural instability improvement in patients with NPH

Shunted NPH patients improved their gait velocity from baseline compared to post-LDT (Supplemental Table 2, *p*<0.001; 95% CI [0.14, 0.23]). Consistent with prior literature, large improvements in stride length were also observed (Supplemental Table 2, *p*<0.001, 95% CI [0.12, 0.22]).^3,9^ For shunted patients, gait stride length then plateaued without any statistically significant changes at 6- and 12-month follow-up when compared to post-LDT (Figure 2A). Patients who were not shunted demonstrated no significant change in gait at any time point (Figure 2B). Pull test step length also improved after LDT for patients who were shunted compared to baseline and continued to improve at 6- and 12-month post-operative visits (Supplemental Table 2, Figure 2C, all *p*<0.001, Baseline to Post-LDT 95% CI [1.66, 3.54], Post-LDT to 6-month follow up 95% CI [0.99, 2.85], 6-month follow up to 12-month follow up 95% CI [0.82, 2.61]). There were no statistically significant changes in step length among non-shunted patients at any time point (Figure 2D, Supplemental Table 2).

**Figure 2.**
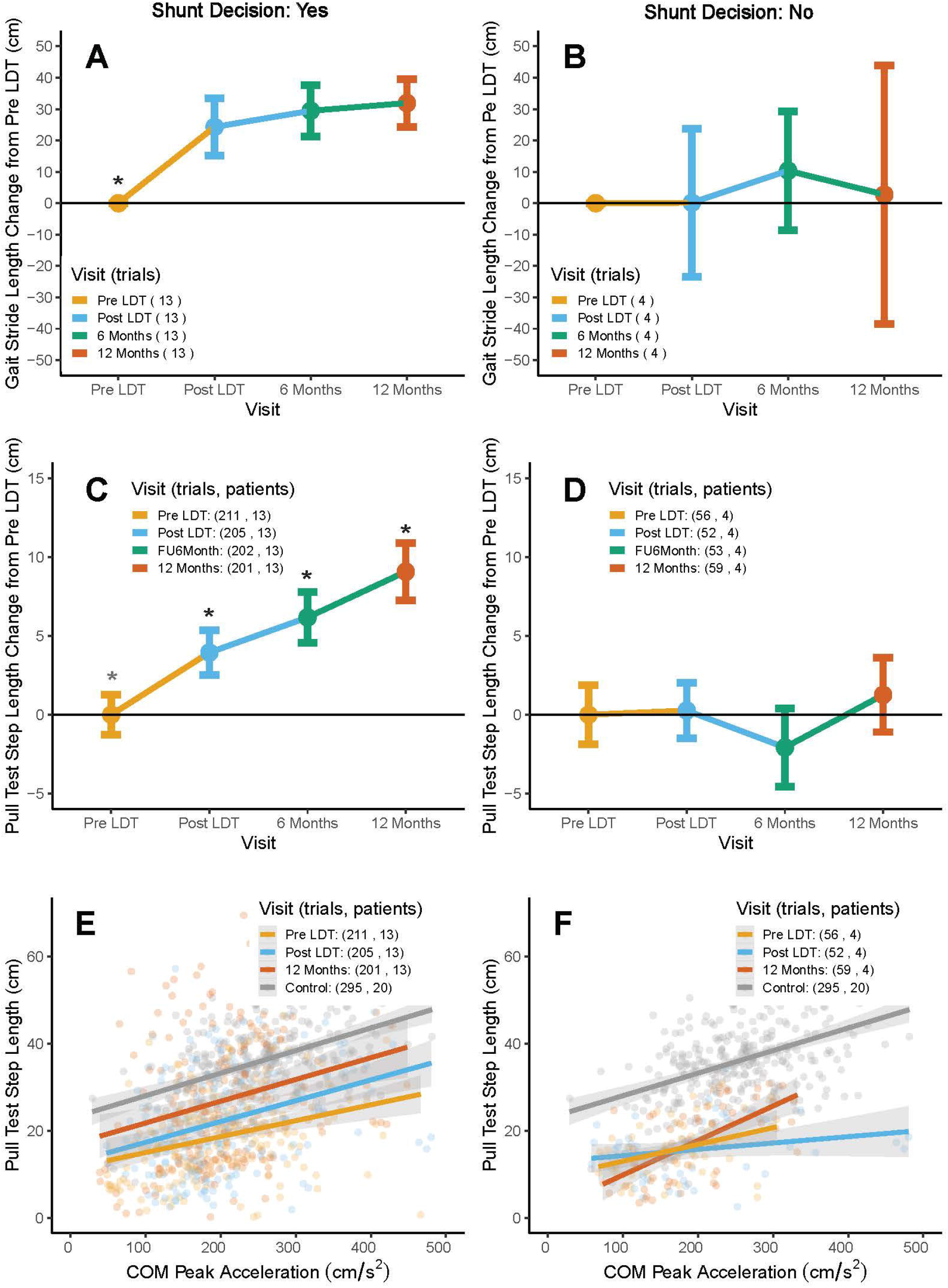

When plotting PTSL against pull intensity, shunted NPH patients visually demonstrated a consistent increase in step length across all pull intensities over time (non-overlapping trend lines with similar slopes) (Figure 2E). Non-shunted patients did not demonstrate any discernible pattern between step length and pull intensity across visits (Figure 2F).

Another component of postural instability, reaction time, demonstrated significant improvement from baseline to 6- and 12-month post-operative follow-ups in shunted patients (Figure 3A, both *p*<0.001, baseline to 6-month follow up 95% CI [-0.10, -0.04], baseline to 12-month 95% CI [95%CI: -0.12, -0.07) but did not show a statistically significant improvement in reaction time post-LDT compared to baseline. Non-shunted patients did demonstrate significantly faster reaction times at 6-month follow-up, but this was not sustained as they regressed prior to their 12-month follow-up (Figure 3B). When reaction time was compared to pull intensity (A_COM_) across visits, patients with lower A_COM_ values appeared to exhibit larger improvements in reaction time compared to patients with higher A_COM_ values (Figure 3C). This trend was not present among the non-shunted patients (Figure 3D).

## DISCUSSION

Our findings demonstrate significantly improved gait impairment and PI in shunted NPH patients over one year. However, our current results suggest two distinct periods of recovery as reflected in PI and gait measures with two important findings that have both theoretical and practical implications. First, gait and postural improvements are both observable after a short-term (3-day) LDT (Figure 2A; 2C), but no further improvements in gait are seen over one year of follow-up, while step length in response to the MDS-UPDRS_PT_ continues to significantly increase throughout the 12 month follow up period (Supplemental Table 2). Second, shunted patients’ overall kinematic outcomes as reflected by their V_COM_ profiles and peak V_COM_ values (Figure 1A) were improved post-LDT, which is most likely due to an increase in step length (Figure 2C), but not reaction time (Figure 3A), despite no change in the clinical MDS-UPDRS_PT_ scores post-LDT (Table 1). This indicates that quantitative kinematic assessment of these patients may be a more sensitive indicator of PI improvement than simply assigning an ordinal score.

**Figure 3.**
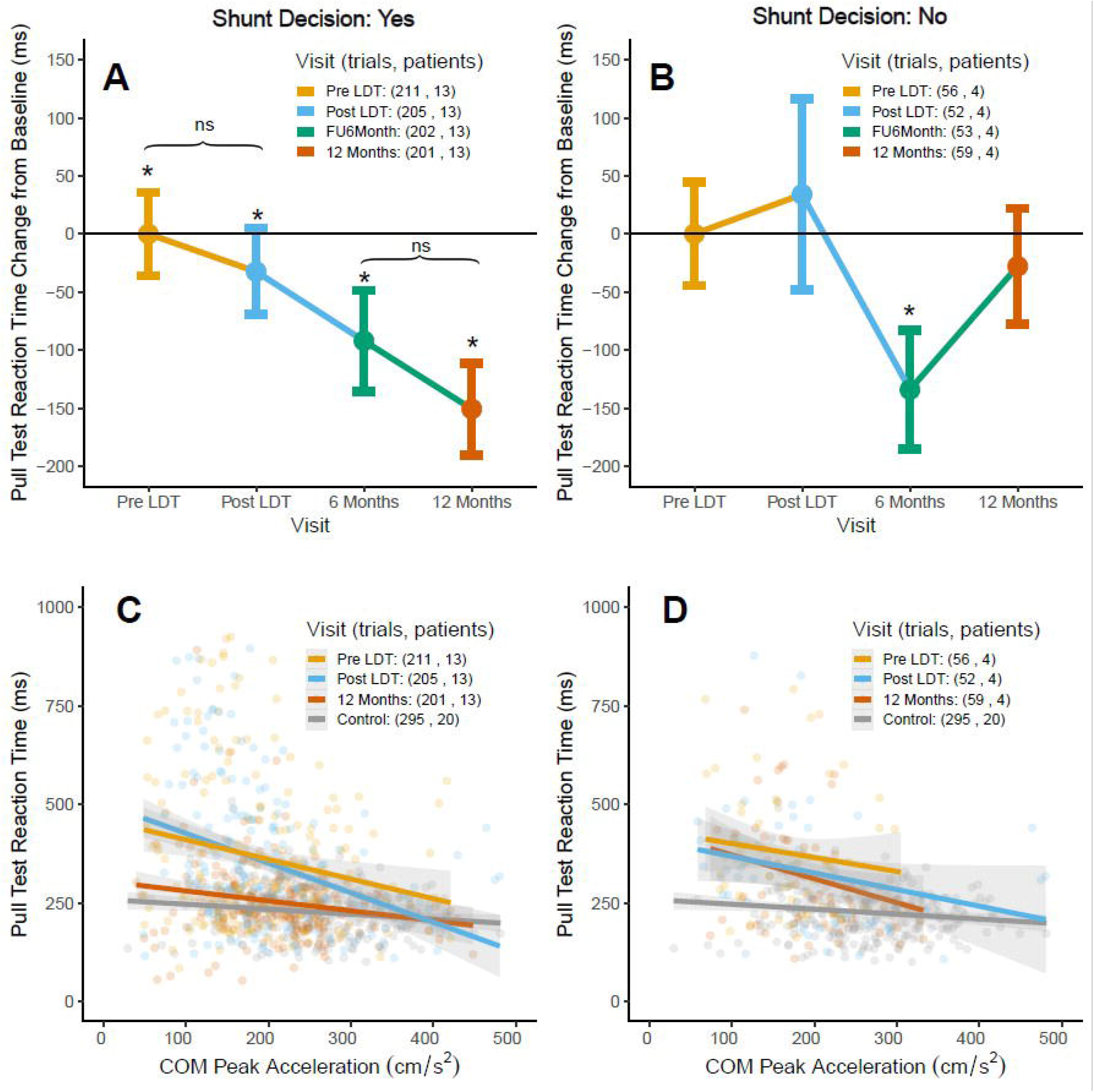

Changes in improvement in gait and PI over time could be the result of differences in measurement, differential responses to changes in valve settings at follow-up visits or differences in physiological mechanisms. First, reactive postural response steps were more variable (by design) and therefore when looking only at mean step length, they may be statistically less distinguishable from visit to visit compared to comfortable gait stride length (Figure 2A,2C). To minimize this limitation, we examined only within-patient changes between visits. We also plotted PTSL across the entire range of pull intensity for each visit and found that patients’ step scaling response (slope) was remarkably consistent across visits while their overall step length (y-intercept) visually increased over the year of follow-up (Figure 2E).

The changes in improvement of postural and gait variables over time could also theoretically represent differences in response to changes in shunt valve settings over time. The treating neurosurgeon changed valve settings at follow-up visits depending on the patient’s clinical status. Typically, this involved dialing the shunt setting down to increase CSF outflow and theoretically improve gait and balance function. If gait responds less to changes in CSF outflow after VPS and PI responds more, this could explain why gait appears to plateau over time while the reactive postural response continues to improve. This implies, however, that there are still fundamentally different physiological mechanisms underlying their improvement.

Most intriguingly, the differentiation over time between gait and PI improvement implies two separate underlying physiological mechanisms. While initially thought of as an “automatic” brainstem-based reflex, reactive postural stability is now recognized to be a complex coordination of visual, vestibular, somatosensory, and proprioceptive inputs that requires cortical and subcortical coordination.^10-12^ The cortical contributions to PI are just beginning to be understood and the improvements seen in shunted NPH patients represent a unique opportunity to investigate the mechanisms of change in future studies via other means such as fMRI and EEG.^13^ This is especially important since other neurodegenerative diseases that feature PI such as Parkinson’s disease have no effective targeted therapies for PI. Investigating EEG patterns of improvement in these patients could lead to targeted neuromodulation-based studies in a variety of movement disorder patients.

The clinical relevance of longitudinal improvements observed in the reactive postural response after VPS is unknown. Our results show a mean increase of 8.6 cm in step length and a decrease of 131 ms in reaction time from baseline (pre-LDT) to 12 months post-operatively for shunted patients. This represents a 46.8% increase in step size when compared to the baseline of 18.4 cm and a 33.2% decrease in reaction time from a baseline of 395 ms. These improvements occurred simultaneously with UPDRS_PT_ score improvements by 12 months (Table 1). We have previously shown that changes in the relationship of the initial step length to perturbation intensity can be related to UPDRS_PT_ score although we have not previously shown differences in reaction time between UPDRS_PT_ scores.^6^ Non-shunted patients did demonstrate an improvement in reaction time at 6 months post-op, but this significance disappeared by one year. This raises the possibility of a learning effect given the number of trials and visits that these patients participate in over the course of a given year. We have previously demonstrated that there are no “within-visit” learning effects as reaction time does not improve with increasing trial number (i.e., patients do not react faster for Trial 15 compared to Trial 1)^6^ but this does not exclude the possibility of long-term learning effects.

Given the magnitude of improvement in step length and its link to UPDRS_PT_ score, we propose this is a main mechanism of clinical improvement in PI in NPH after VPS. Reaction time clearly improves over time as well although how much this contributes to the improvement, whether it is partly due to learning effects and whether this remains part of the same physiological system underlying improvement in PI in NPH remains undetermined. Further research should also be aimed at confirming this in a larger patient sample, relating these changes to patient-reported outcome measures, and creating minimal clinically important differences for these kinematic parameters. This will better define the relationship of kinematics to clinician and patient-reported outcomes to establish the clinical relevance of these changes after VPS.

No prior studies have examined whether PI improves in patients with NPH longitudinally after VPS. The observed pattern of improvement implies that there may be linear, long-term changes that occur due to CSF diversion via VPS. This timescale of recovery is significantly longer than that seen in gait velocity and stride length and therefore further investigation utilizing EEG, and/or fMRI in combination with kinematics is warranted to explore the neurological pathophysiology leading to these differences.

## Data Availability

All data produced in the present study are available upon reasonable request to the authors

## Supplementary material

Supplementary material is available at *Journal of Neurology, Neurosurgery & Psychiatry* online.

## References

1. Adams, R., Fisher, C., Hakim, S., Ojemann, R., Sweet, W. Symptomatic occult hydrocephalus with “normal” cerebrospinal fluid pressure. A treatable syndrome. N Engl J Med. 1965 Jul 15;273:117–26. doi:10.1056/NEJM196507152730301

2. Davis A, Luciano M, Moghekar A, Yasar S. Assessing the predictive value of common gait measure for predicting falls in patients presenting with suspected normal pressure hydrocephalus. BMC Neurol. 2021;21(1):60. Published 2021 Feb 8. doi:10.1186/s12883-021-02068-0

3. Stolze H, Kuhtz-Buschbeck JP, Drücke H, Jöhnk K, Illert M, Deuschl G. Comparative analysis of the gait disorder of normal pressure hydrocephalus and Parkinson’s disease. J Neurol Neurosurg Psychiatry. 2001;70(3):289–297. doi:10.1136/jnnp.70.3.289

4. Pujari S, Kharkar S, Metellus P, Shuck J, Williams MA, Rigamonti D. Normal pressure hydrocephalus: long-term outcome after shunt surgery. J Neurol Neurosurg Psychiatry. 2008;79(11):1282–1286. doi:10.1136/jnnp.2007.123620

5. Grasso G, Torregrossa F, Leone L, Frisella A, Landi A. Long-Term Efficacy of Shunt Therapy in Idiopathic Normal Pressure Hydrocephalus. World Neurosurg. 2019;129:e458–e463. doi:10.1016/j.wneu.2019.05.183

6. Daly S, Hanson JT, Mavanji V, et al. Using kinematics to re-define the pull test as a quantitative biomarker of the postural response in normal pressure hydrocephalus patients. Exp Brain Res. 2022;240(3):791–802. doi:10.1007/s00221-021-06292-5

7. Goetz CG, Tilley BC, Shaftman SR, et al. Movement Disorder Society-sponsored revision of the Unified Parkinson’s Disease Rating Scale (MDS-UPDRS): scale presentation and clinimetric testing results. Mov Disord. 2008;23(15):2129–2170. doi:10.1002/mds.22340

8. Schepers M, Giuberti M, Bellusci G. Xsens MVN: Consistent Tracking of Human Motion Using Inertial Sensing. Xsens Technologies. March 2018:1–8. doi:10.13140/RG.2.2.22099.07205

9. Bugalho P, Guimarães J. Gait disturbance in normal pressure hydrocephalus: a clinical study. Parkinsonism Relat Disord. 2007;13(7):434–437. doi:10.1016/j.parkreldis.2006.08.007

10. Liddell E, Sherrington C. Reflexes in Response to Stretch (Myotatic Reflexes). Proceedings of the Royal Society of London Series BContaining Papers of a Biological Character. 1924;96(675):212–242

11. Lord SR, Menz HB. Visual contributions to postural stability in older adults. Gerontology. 2000;46(6):306–310. doi:10.1159/000022182

12. Takakusaki K. Functional Neuroanatomy for Posture and Gait Control. J Mov Disord. 2017;10(1):1–17. doi:10.14802/jmd.16062

13. Palmer JA, Payne AM, Ting LH, Borich MR. Cortical Engagement Metrics During Reactive Balance Are Associated With Distinct Aspects of Balance Behavior in Older Adults. Front Aging Neurosci. 2021;13:684743. Published 2021 Jul 14. doi:10.3389/fnagi.2021.684743

