## Supplementary figures and images for "Longitudinal improvement in postural instability is distinct from gait in treated adult hydrocephalus"

### Supplemental Figure 1

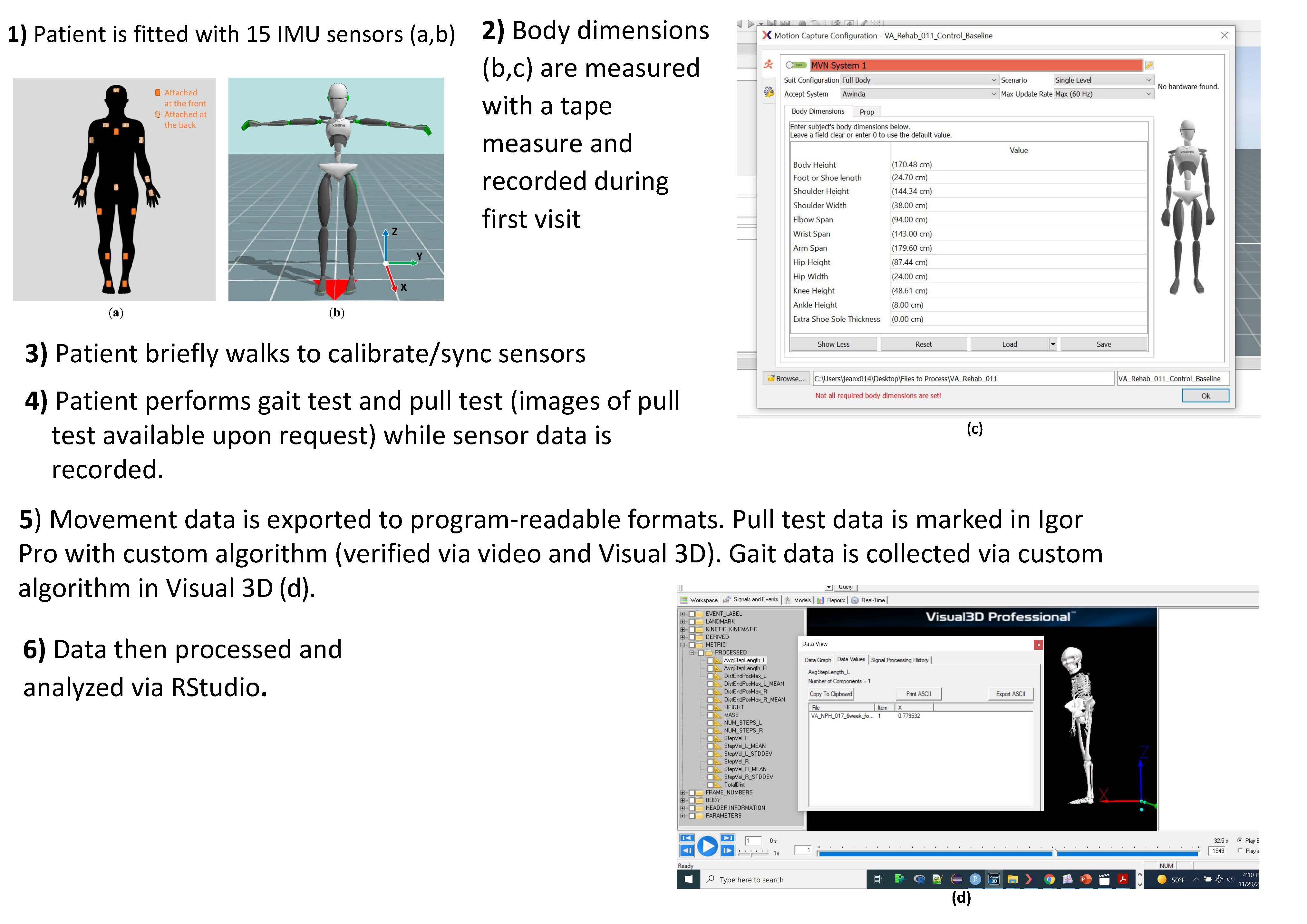

### Supplemental Figure 2

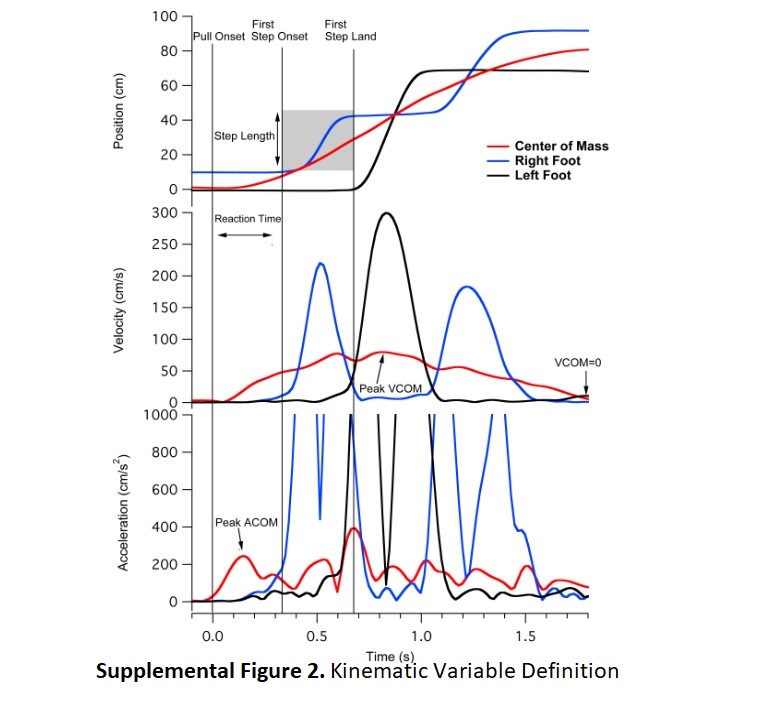

### Supplemental Figure 3

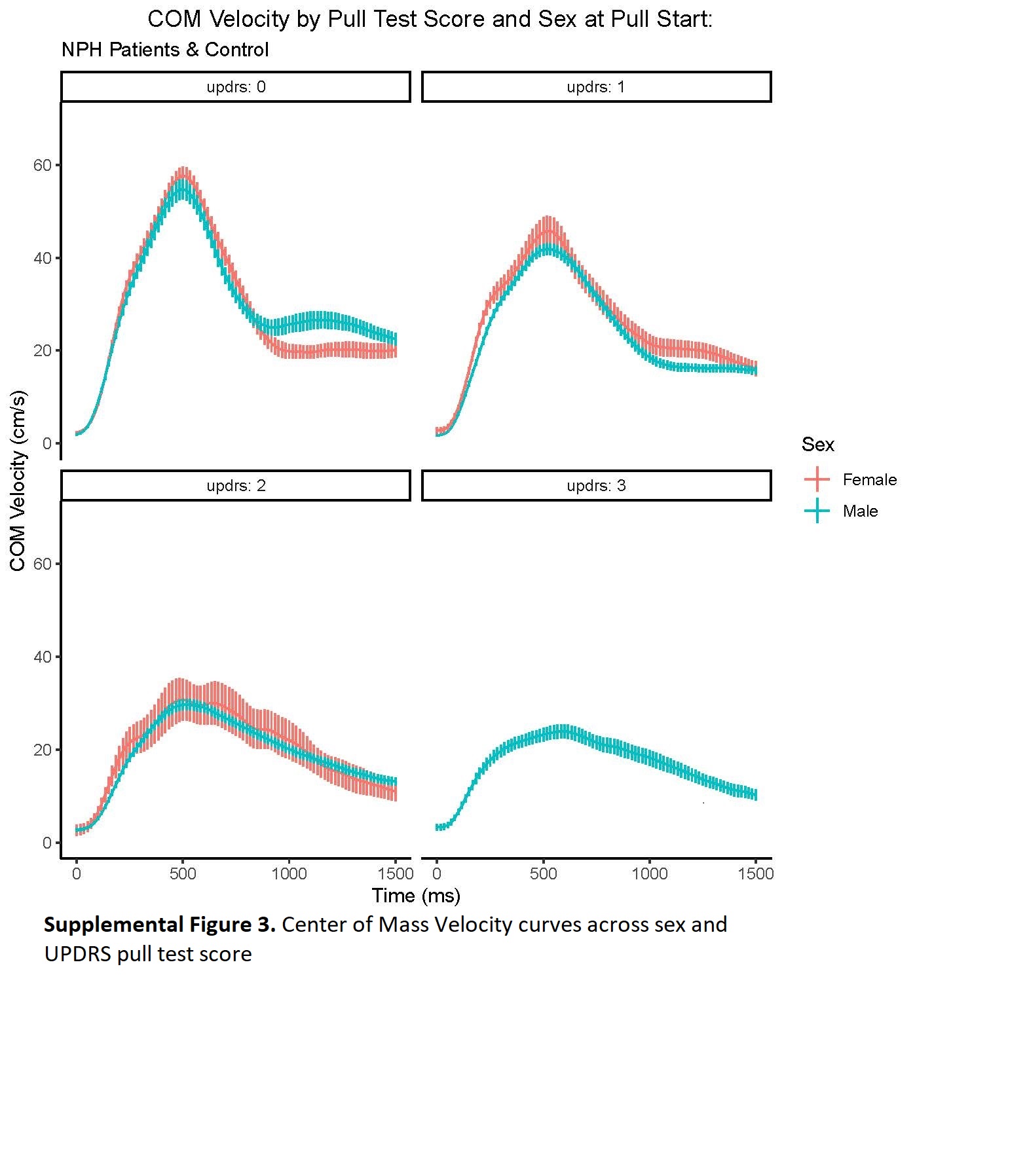
