## Supplemental Table 1 for "Longitudinal improvement in postural instability is distinct from gait in treated adult hydrocephalus"

| **Demographic:** | **Shunt Decision** | | |
| --- | --- | --- | --- |
|  | ***Yes*** | ***No*** | ***Control*** |
| **Sex** | *11 M , 2 F* | *4 M , 0 F* | *4 M, 16 F* |
| **Weight in kg (SD)** | *96.25 (20.99)* | *78.88 (21.86)* | *70.24 (11.99)* |
| **Age in years (SD)** | *71.92 (7.17)* | *71.75 (6.29)* | *70.00 (4.00)* |
| **Height in cm (SD)** | *174.75 (9.34)* | *170.08 (9.03)* | *169.45 (9.53)* |

***Supplemental Table 1 Patient Demographics***
