## Supplemental Table 2 for "Longitudinal improvement in postural instability is distinct from gait in treated adult hydrocephalus"

.Supplemental Table 2: Assessing absolute changes over time.

| Shunted | Outcome | Change from Baseline to Post-LDT | Change from Baseline to  6-month | Change from Baseline to 12-month | Change from Post-LDT to 6-month | Change from Post-LDT to 12-month | Change from 6-month to 12-month |
| --- | --- | --- | --- | --- | --- | --- | --- |
| Yes | Peak COM Velocity (cm/s)^1^ | **4.67 (95%CI: 3.16, 6.18); p<0.001** | **5.07 (95%CI: 3.53, 6.60); p<0.001** | **9.02 (95%CI: 7.18, 10.86); p<0.001** | 1.20 (95%CI: -0.16, 2.57); p=0.836 | **4.50 (95%CI: 2.80, 6.20); p<0.001** | **3.39 (95% CI:1.89,4.93 ): p<0.001** |
|  | Time to Peak Velocity (s)^1^ | -0.03 (95%CI: -0.08, 0.02); p>0.999 | **-0.07 (95%CI: -0.12, -0.03); p=0.027** | **-0.08 (95%CI: -0.12, -0.04); p=0.002** | -0.06 (95%CI: -0.11, -0.01); p=0.269 | -0.06 (95%CI: -0.10, -0.01); p=0.100 | 0.002 (95% CI:  -0.04, 0.04) p>0.999 |
|  | Gait Velocity (m/s)^2^ | **0.19 (95%CI: 0.14, 0.23); p<0.001** | **0.19 (95%CI: 0.13, 0.25); p<0.001** | **0.25 (95%CI: 0.19, 0.31); p<0.001** | >0.001 (95%CI:  -0.08, 0.08); p>0.999 | 0.06 (95%CI:  -0.01,0.01); p = 0.63 | 0.06 (95% CI: -.02, 0.15)  p = 0.859 |
|  | Stride Length (m)^2^ | **0.17 (95%CI: 0.12, 0.22); p<0.001** | **0.21 (95%CI: 0.16, 0.26); p<0.001** | **0.23 (95%CI: 0.18, 0.27); p<0.001** | 0.04 (95%CI: -0.04, 0.11); p>0.999 | 0.05 (95%CI: -0.02, 0.13); p=0.850 | -.0.01 (95% CI: -0.09, 0.05) p>0.999 |
|  | Pull Step Length (cm)^1^ | **2.60 (95%CI: 1.66, 3.54); p<0.001** | **4.01 (95%CI: 2.98, 5.03); p<0.001** | **6.08 (95%CI: 4.99, 7.17); p<0.001** | **1.92 (95%CI: 0.99, 2.85); p<0.001** | **3.60 (95%CI: 2.63, 4.58); p<0.001** | **1.72 (95% CI: 0.82,2.62) p<0.001** |
|  | Pull Reaction Time (s)^1^ | -0.03 (95%CI: -0.05, 0.00); p=0.367 | **-0.07 (95%CI: -0.10, -0.04); p<0.001** | **-0.10 (95%CI: -0.12, -0.07); p<0.001** | **-0.05 (95%CI: -0.08, -0.02); p=0.007** | **-0.08 (95%CI: -0.10, -0.05); p<0.001** | 0.06 (95% CI:  -0.05, 0.00) p=0.333 |
| No | Peak COM Velocity (cm/s)^1^ | -1.36 (95%CI: -3.86, 1.15); p>0.999 | -1.19 (95%CI: -3.64, 1.25); p>0.999 | -0.27 (95%CI: -2.63, 2.09); p>0.999 | 1.31 (95%CI: -1.08, 3.70); p>0.999 | 2.69 (95%CI: 0.49, 4.88); p=0.164 | 0.53 (95% CI  -1.62, 2.69) p>0.999 |
|  | Time to Peak Velocity (s)^1^ | 0.06 (95%CI: -0.04, 0.16); p>0.999 | -0.07 (95%CI: -0.12, -0.02); p=0.077 | **-0.06 (95%CI: -0.11, -0.01); p=0.002** | -0.13 (95%CI: -0.23, -0.03); p=0.117 | -0.12 (95%CI: -0.22, -0.03); p=0.090 | 0.01 (95% CI:  -0.05, 0.07)  p>0.999 |
|  | Gait Velocity (m/s)^2^ | -0.01 (95%CI: -0.09, 0.07); p>0.999 | 0.09 (95%CI: -0.03, 0.20); p>0.768 | 0.02 (95%CI: -0.17, 0.21); p>0.999 | 0.10 (95%CI: -0.04, 0.24); p>0.999 | 0.03 (95%CI: -0.15, 0.21); p>0.999 | -0.07 (95% CI: -.29, 0.15)  p =>0.999 |
|  | Stride Length (m)^2^ | 0.00 (95%CI: -0.10, 0.10); p>0.999 | 0.07 (95%CI: -0.01, 0.16); p=0.610 | 0.02 (95%CI: -0.15, 0.19); p>0.999 | 0.07 (95%CI: -0.06, 0.20); p>0.999 | 0.02 (95%CI: -0.16, 0.20); p<0.999 | 0.05 (95% CI:  -0.13, 0.24) p>0.999 |
|  | Pull Step Length (cm)^1^ | -0.35 (95%CI: -1.82, 1.11); p>0.999 | -1.09 (95%CI: -2.56, 0.37); p=0.864 | -0.03 (95%CI: -1.46, 1.40); p>0.999 | -0.49 (95%CI: -1.96, 0.98); p>0.999 | 1.36 (95%CI: 0.00, 2.72); p=0.303 | 0.62 (95% CI:  -0.82, 2.05) p>0.999 |
|  | Pull Reaction Time (s)^1^ | 0.02 (95%CI: -0.04, 0.07); p>0.999 | **-0.09 (95%CI: -0.12, -0.06); p<0.001** | -0.03 (95%CI: -0.07, 0.00); p=0.347 | **-0.11 (95%CI: -0.17, -0.06); p=0.001** | -0.06 (95%CI: -0.11, -0.01); p=0.174 | **0.05 (95% CI: 0.02, 0.09) p=0.003** |

^1^Modeled by mixed-effects regression with random intercept for patient.

^2^Modeled by linear regression.

All models adjusted for baseline value.

p-value includes a Bonferroni correction to account for multiple testing.
